# Association between City-wide Lockdown and COVID-19 Hospitalization Rates in Multigenerational Households in New York City

**DOI:** 10.1101/2021.08.31.21262914

**Authors:** Arnab K. Ghosh, Sara Venkatraman, Evgeniya Reshetnyak, Mangala Rajan, Anjile An, John K. Chae, Mark A. Unruh, David Abramson, Charles DiMaggio, Nathaniel Hupert

## Abstract

**Background:** City-wide lockdowns and school closures have demonstrably impacted COVID-19 transmission. However, simulation studies have suggested an increased risk of COVID-19 related morbidity for older individuals inoculated by house-bound children. This study examines whether the March 2020 lockdown in New York City (NYC) was associated with higher COVID-19 hospitalization rates in neighborhoods with larger proportions of multigenerational households.

**Methods:** We obtained daily age-segmented COVID-19 hospitalization counts in each of 166 ZIP code tabulation areas (ZCTAs) in NYC. Using Bayesian Poisson regression models that account for spatiotemporal dependencies between ZCTAs, as well as socioeconomic risk factors, we conducted a difference-in-differences study amongst ZCTA-level hospitalization rates from February 23 to May 2, 2020. We compared ZCTAs in the lowest quartile of multigenerational housing to other quartiles before and after the lockdown.

**Findings:** Among individuals over 55 years, the lockdown was associated with higher COVID-19 hospitalization rates in ZCTAs with more multigenerational households. The greatest difference occurred three weeks after lockdown: Q2 vs. Q1: 54% increase (95% Bayesian credible intervals: 22 – 96%); Q3 vs. Q1: 48%, (17 – 89%); Q4 vs. Q1: 66%, (30 – 211%). After accounting for pandemic-related population shifts, a significant difference was observed only in Q4 ZCTAs: 37% (7 –76%).

**Interpretation:** By increasing house-bound mixing across older and younger age groups, city-wide lockdown mandates imposed during the growth of COVID-19 cases may have inadvertently, but transiently, contributed to increased transmission in multigenerational households.

**Funding:** National Center for Advancing Translational Sciences; Clinical and Translational Science Center at Weill Cornell Medical College.

## Introduction

Since COVID-19’s first appearance in December 2019, knowledge of transmission dynamics of SARS-CoV2 have rapidly advanced. Studies show that both proximity to infected patients and the concentration of inoculum play important roles in acquisition and subsequent severity of COVID-19. ^1-5^ For this reason, public health measures limiting physical proximity (e.g., social distancing and lockdowns) have been critical to decreasing COVID-19 transmission, and subsequent morbidity and mortality in the United States and abroad.^6^

Evidence suggests that the organization of households and housing structure may also play a role in COVID-19 transmission. Ecological studies that examined links between housing conditions and COVID-19 rates describe associations between poorer housing conditions and COVID-19 cases,^7^ and between overcrowded housing and COVID-19 test positivity,^8-11^ even after accounting for numerous socioeconomic factors. Furthermore, multigenerational households may be at further risk because of the skewed nature of COVID-19 morbidity towards older individuals.^12,13^ In a nationwide study of multigenerational households in the United Kingdom during their first wave, men and women living in multigenerational households with children had a 17-21% increased risk of COVID- 19 mortality after adjusting for several socioeconomic and clinical risk factors.^14^

Non-pharmaceutical interventions (NPI) such as city-wide lockdowns and school closures have had demonstrable impacts on COVID-19 transmission.^15-17^ However, results from simulation modeling studies have suggested an increased risk of COVID-19 related morbidity after NPIs including school closures, driven by increased inoculation of older individuals by house-bound school-age children, ^18^ particularly in communities with limited means to reduce other potential contacts.^19^ This hypothesis has not been empirically tested using real world data. To explore this, we investigated whether the New York City (NYC)-wide lockdown in spring 2020 was associated with increased COVID-19 hospitalization rates in ZIP codes with higher proportions of multigenerational households. We hypothesized that after lockdown, COVID-19 hospitalization rates among older residents would be disproportionately higher in those ZIP codes driven by high-risk older individuals contracting COVID-19 from asymptomatic school-aged children by sharing the same indoor space in households.

## Methods

### Study Setting

We conducted a retrospective ZCTA-level difference-in-difference analysis of weekly COVID- 19 hospitalizations per 10,000 population in each ZCTA in NYC between February 23 and May 23, 2020. Beginning in late February 2020, NYC became the global epicenter of the COVID-19 pandemic, with a first wave that crested in late March/early April 2020 (Figure 1). A city-mandated closures of all schools starting March 16, 2020 was the first public health measure to address the growing tide of COVID-19 cases,^20^ followed by a state-wide lockdown four days later.^21^

**Figure 1:**
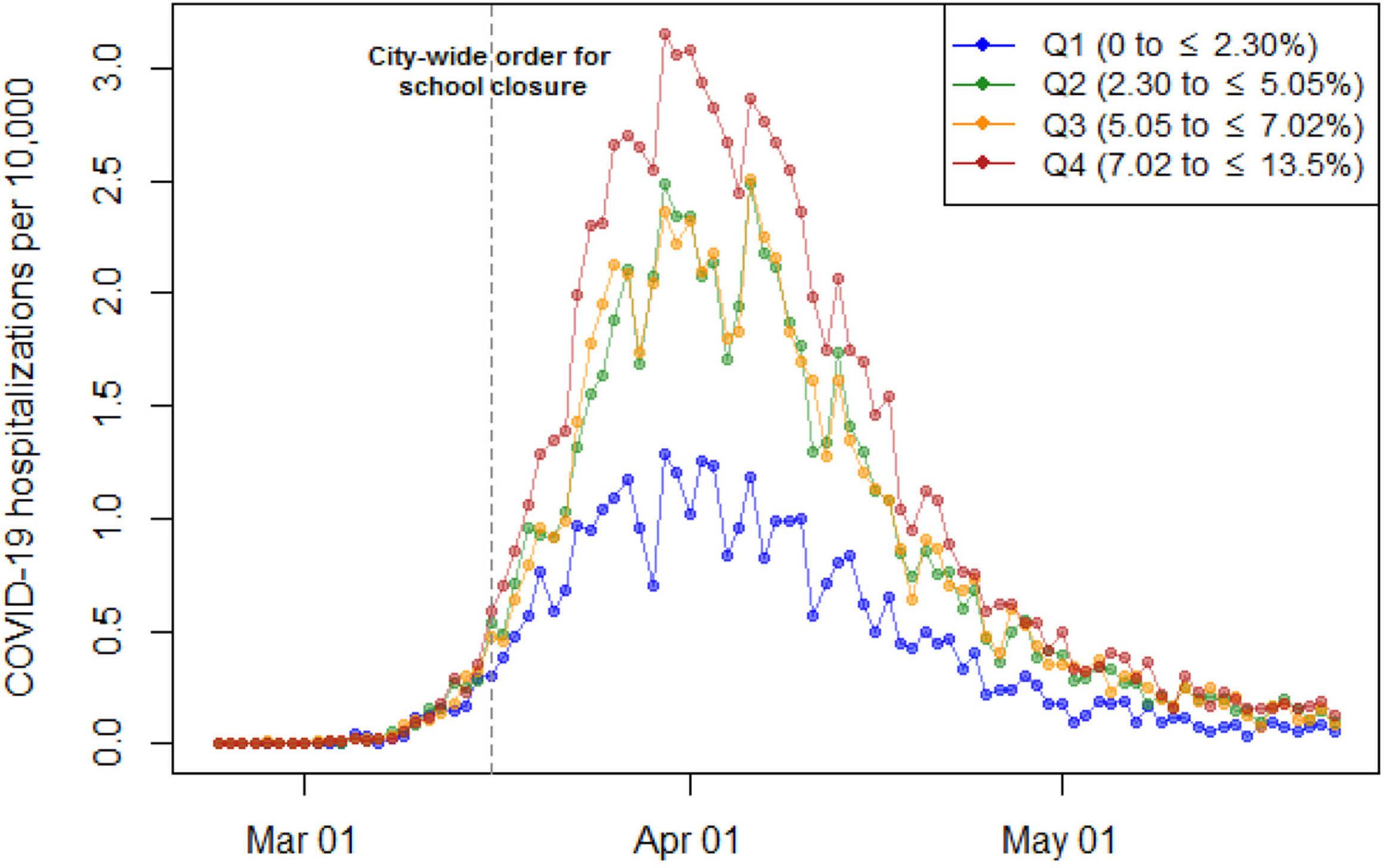
Unadjusted trends in COVID19 hospitalizations per 10,000 in New York City (NYC) by quartiles of multigenerational households, February 23 to May 23, 2020.

### Variables and Data Sources

Our dependent variable was weekly COVID-19 hospitalization counts which were used to calculate rates per 10,000 population on all patients and those over 55 years of age by ZCTA, aggregated from daily COVID-19 hospitalization counts obtained from the NYC Department of Health and Mental Hygiene (NYC DOHMH). We calculated the proportion of multigenerational households (defined as the percentage of households occupied by a grandparent and a grandchild less than 18 years of age) within each ZCTA using the American Community Survey 5-year 2018 estimates. The primary analysis focused on hospitalized patients aged 55 years and older to reflect the average age of grandparents in the US.^22^ Secondary analyses examined COVID-19 related hospitalized patients of all ages.

Although several socioeconomic factors have been reported to be associated with increased rates of COVID-19 cases, we were careful to consider the potential mediating effect of these factors in order to prevent overadjusting our model. In order to account for socioeconomic factors that likely confound the ecological-level association between the lockdown, COVID-19 hospitalization rates in ZCTAs, and multigenerational households, we modified the conceptual model used by Nalfilyan et al.;^14^ this study described the relationship between household composition, socioeconomic status, and COVID-19 related risk.

Therefore, our primary analysis included, by ZCTA, the proportion of White residents, proportion of residents living under the federal poverty line (both as a percentage) and the median income (in US dollars, 2018). Furthermore, in prior work on the first COVID-19 wave in NYC,^23^ we also included the proportion of overcrowded households in the model as a control covariate, where overcrowdedness was defined as estimated number of housing units with more than one occupant per room, divided by the number of occupied housing units, expressed as a percentage. All measures of these covariates were taken from the American Community Survey (ACS) 5-year 2018 estimates.

In order to undertake inferential spatial analyses, spatial shapefiles of NYC’s ZCTAs were downloaded from the New York City Department of City Planning.

### Outcome

The primary outcome of interest was the adjusted COVID-19 weekly hospitalization rate per 10,000 population within each ZCTA for a) hospitalized patients over 55 years old, and b) for all hospitalized patients. We calculated these rates by ZCTA quartiles of the multigenerational households using the first (lowest) quartile as the reference group.

### Statistical Analysis

We first compared the overall number of COVID-19 hospitalizations, COVID-19 hospitalizations per 10,000, and the socioeconomic characteristics of ZCTAs by quartile of multigenerational households. Visual depictions of the COVID-19 hospitalization rates per 10,000 were created to describe the trajectory of the COVID-19 pandemic, segmented by each quartile of multigenerational households.

We then estimated the association between the city-wide lockdown that began with lockdown and the COVID-19 hospitalization rates within each quartile of multigenerational households, using a modified difference-in-difference analysis. Whereas a typical difference-in-difference analysis defines an unexposed control group, and an exposed treatment group (in our analysis, the exposure is the city-wide lockdown beginning with school closures that began on March 16, 2020), our modified difference-in-difference analysis hypothesized an effect of increasing magnitude on COVID-19 hospitalization rates for ZCTAs with higher proportions of multigenerational households (quartiles 2-4) in relation to quartile 1. Therefore, for the purpose of this analysis, we treated ZCTAs in the first quartile of multigenerational households as the control group.

We could not test the parallel trends assumption of the difference-in-difference methodology directly. However, we assessed trends in the daily and weekly COVID-19 hospitalization rates by quartile of multigenerational households prior to lockdown. Furthermore, we believe the assumption holds as the decision for the lockdown was unlikely to be associated with other factors since it was abruptly implemented at the same time across all ZCTAs.

In our analysis, we estimated multivariable generalized linear models specifying a Poisson distribution for the weekly COVID-19 hospitalization rates. In these models, a binary indicator was included for denoted each week before and after the city-wide lockdown order. This allowed us to compare the COVID-19 hospitalization rates for each quartile of multigenerational households relative to the first quartile, in weekly increments before and after the lockdown took effect. Given the known clustering of COVID-19 cases in NYC, we used Moran’s I to formally assess spatial autocorrelation of the cumulative COVID-19 hospitalizations rates by ZCTA over the study period. Details of our model specification are provided in the Supplement (eFigure 2).

**Figure 2:**
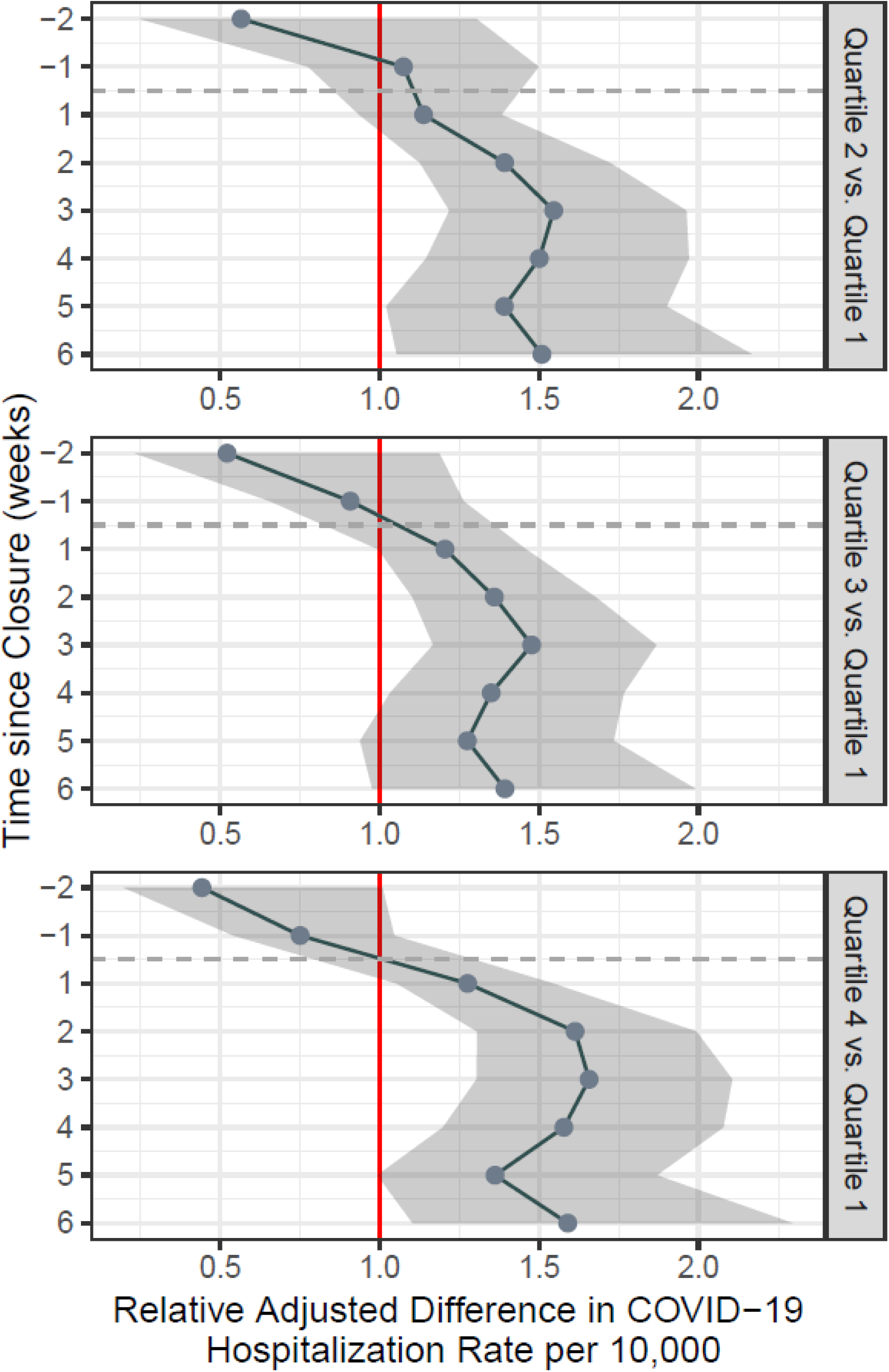
Adjusted Difference-in-Differences Estimates of the Association Between School Closure and Adjusted COVID-19 Hospitalization Rates for Patients Older than 55 years by Quartiles of Multigenerational ZIP Codes, with Quartile 1 as reference.^1,2^. 1 In all panels, the Y-axis represents the number of weeks relative to the school closure order, with event weeks 2 weeks before the exposure, and up to 6 weeks after the exposure 2 Adjusted for the following ZCTA-level covariates: percentage of overcrowded households (defined as estimated number of housing units with more than one occupant per room, divided by the number of occupied housing units), percentage of White residents, percentage of residents living under the Federal Poverty Line, and median income in USD, 2018 – all taken from the American Community Survey 2018 5-year estimates.

Upon finding that there was significant spatial autocorrelation in our dependent variable (Moran’s I: 0.575, p = 0.001 – Supplement), our final model was a Bayesian version of a Poisson regression model that we fit using the integrated nested Laplace approximation (INLA) method. The INLA is a computationally efficient method for fitting models to data exhibiting spatial or temporal structure, and has been used in other ecological analyses of COVID-19.^10,24^ Coefficients, derived from the multivariable INLA model and their 95% Bayesian credible intervals, are presented. These coefficients represent the percent increased risk for COVID-19 hospitalizations in ZCTAs with higher multigenerational housing (quartiles 2-4) compared to quartile 1. We then fit a linear regression model to panel data over all age groups for each ZCTA.

### Secondary Analyses

We performed two subsequent analyses. The first modelled all COVID-19 hospitalizations regardless of age group using the same covariates as the primary analysis. The model used in this analysis includes an offset term accounting for ZCTA-level population size.

The second analysis employed the same analytical model as the primary analysis but also accounted for population shifts related to pandemic-related flight. During the first COVID-19 wave, GPS data suggested that large segments of the population left NYC with residents from wealthier neighborhoods leaving in larger proportions.^25^ We adjusted ZCTA-level population estimates according to census tract-level population shifts taken from publicly available cellular device data from Teralytics between January 1, 2020 and April 15, 2020, obtained from the New York Times.^26^ Informed by different data from Descartes Labs,^25^ we modeled population shifts resulting from pandemic-related flight as time-varying, reaching a stable population estimate two weeks after lockdown.

### Sensitivity Analyses

We conducted two sensitivity analyses to test the robustness of our findings (Supplement). To test whether our results could be explained by other potential confounding socioeconomic characteristics, we expanded the number of ZCTA-level control covariates to include the proportion of essential workers who were more likely to work in the community and not at home (calculated according to the methodology described elsewhere),^23,27^ the prevalence of COVID-19 related clinical risk factors such as obesity, diabetes, coronary heart disease, smoking and chronic obstructive airways disease. Second, we modeled the proportion of overcrowded households (defined above) jointly with quartiles of multigenerational households to test the robustness of the hypothesized relationship between multigenerational households and COVID-19 hospitalization after accounting for the spatiotemporal relationship of overcrowded ZCTAs and the city-wide lockdown. Because age-specific area-level chronic disease prevalence statistics are not available, these sensitivity analyses were conducted using COVID-19 hospitalization data for all ages.

The study protocol was approved by Weill Cornell Medical College Institutional Review Board. All analyses were conducted in GeoDa version 1.16, QGIS version 3.16.2, and R version 3.6.2. Data were analyzed between January 20 and May 25, 2021.

## Results

### Unadjusted Estimates

During the study period, there were 51,008 individuals hospitalized with COVID-19 across 166 ZCTAs in NYC, 71.05% of which were patients over 55 years. As the quartile of multigenerational households moved from first to fourth quartile (i.e., from smallest to largest proportion of multigenerational households), the total number of hospitalized COVID-19 cases in each quartile increased overall, as well as per 10,000 population (Table 1). Compared to ZCTAs in the first quartile, the number of COVID-19 hospitalizations was more than two-fold higher (Q1: 33.95 hospitalizations per 10,000 vs. Q4: 82.21 hospitalizations per 10,000).

**Table 1:**
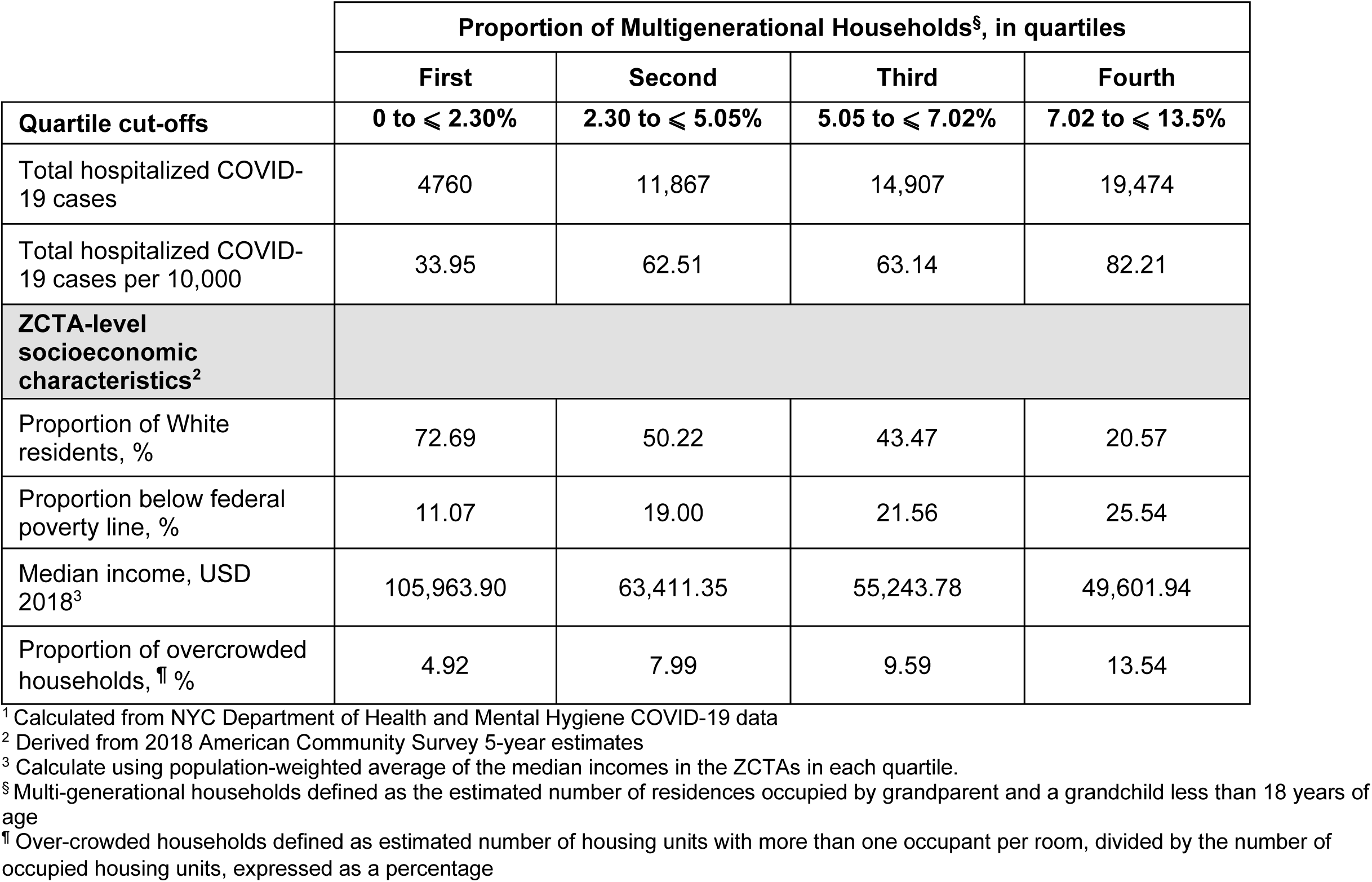
New York City (NYC) ZIP code tabulation area (ZCTA)-level COVID-19 hospitalization cases,^1^ rates, and socioeconomic characteristics by proportion of multigenerational household, in quartiles, February 23 to May 2, 2020.

As ZCTAs contained more multigenerational households, the median income more than halved (Q1: $105,975.40 vs. Q4: $49,647.22), the proportion of White residents fell by more than a third (Q1: 72.49% vs. Q4: 20.68%), the proportion of residents below the federal poverty line approximately doubled (Q1: 11.01% vs. Q4: 25.62%), and the proportion of those residing in overcrowded housing increased (Q1: 4.92% vs. Q4: 13.53%).

Figure 1 displays the unadjusted daily trend of COVID-19 hospitalizations in ZCTAs in each quartile of multigenerational households. Figure 1 is extended over the study period to May 23, 2020 to illustrate the down-trending daily COVID-19 hospitalization rate. The peak of COVID-19 hospitalizations in NYC took place on March 30, 2020. At the peak of hospitalizations, there is almost a three-fold difference in unadjusted COVID-19 hospitalization counts between quartiles (March 30, 2020; Quartile 1: 1.28 cases per 10,000 vs. Quartile 4: 3.16 cases per 10,000).

### Adjusted Estimates

Figure 2 shows estimates of the relative adjusted differences in COVID-19 hospitalization rates for patients over 55 years of age, as compared to Quartile 1 with respect to the city-wide lockdown. Prior to the lockdown’s introduction, there were no significant differences in COVID-19 hospitalization rates relative to quartile 1 (i.e., ZCTAs with the lowest proportion of multigenerational households). After lockdown, there was a consistent and significant increase in COVID-19 hospitalization rates across all ZCTA quartiles. This difference persists over the study period for all quartiles. The greatest difference in adjusted COVID-19 hospitalization rates for all quartiles in reference to quartile 1 was noted 3 weeks after lockdown, Q2 vs. Q1: 1.55 (95% CI: 1.22 – 1.96); Q3 vs. Q1: 1.48, (95% CI: 1.17 – 1.87); Q4 vs. Q1: 1.66, (95% CI: 1.30 – 2.10). These findings suggest that at 3 weeks after lockdown, there was between a 55-66% increased risk of hospitalization from COVID-19 in ZCTAs with higher proportions of multigenerational households compared to ZCTAs with the lowest proportion of multigenerational households.

### Secondary Analysis

Secondary analyses of all patients hospitalized with COVID-19 demonstrated similar findings to the primary analysis (Figure 3A). Relative differences between adjusted COVID-19 hospitalization rates in the first quartile and second, third, and fourth quartiles were greatest at week 3; Q2 vs. Q1: 1.62 (95% CI: 1.33 – 2.00); Q3 vs. Q1: 1.60, (95% CI: 1.32 – 1.96); Q4 vs. Q1: 1.62, (95% CI: 1.32 – 1.99). However, the pre-closure adjusted COVID-19 hospitalization rate was higher in the first quartile compared to the fourth quartile, and the difference was statistically significant.

**Figure 3:**
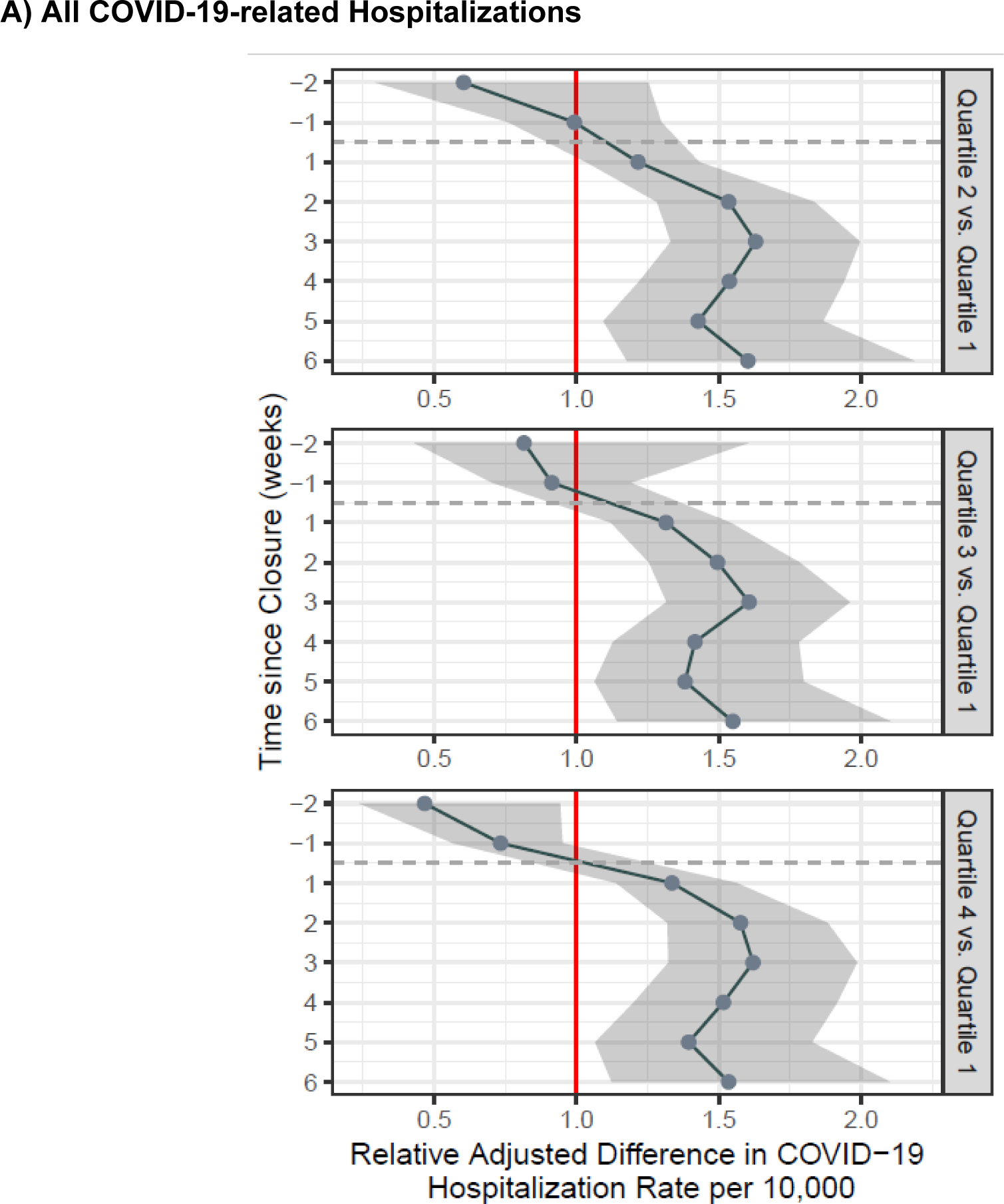
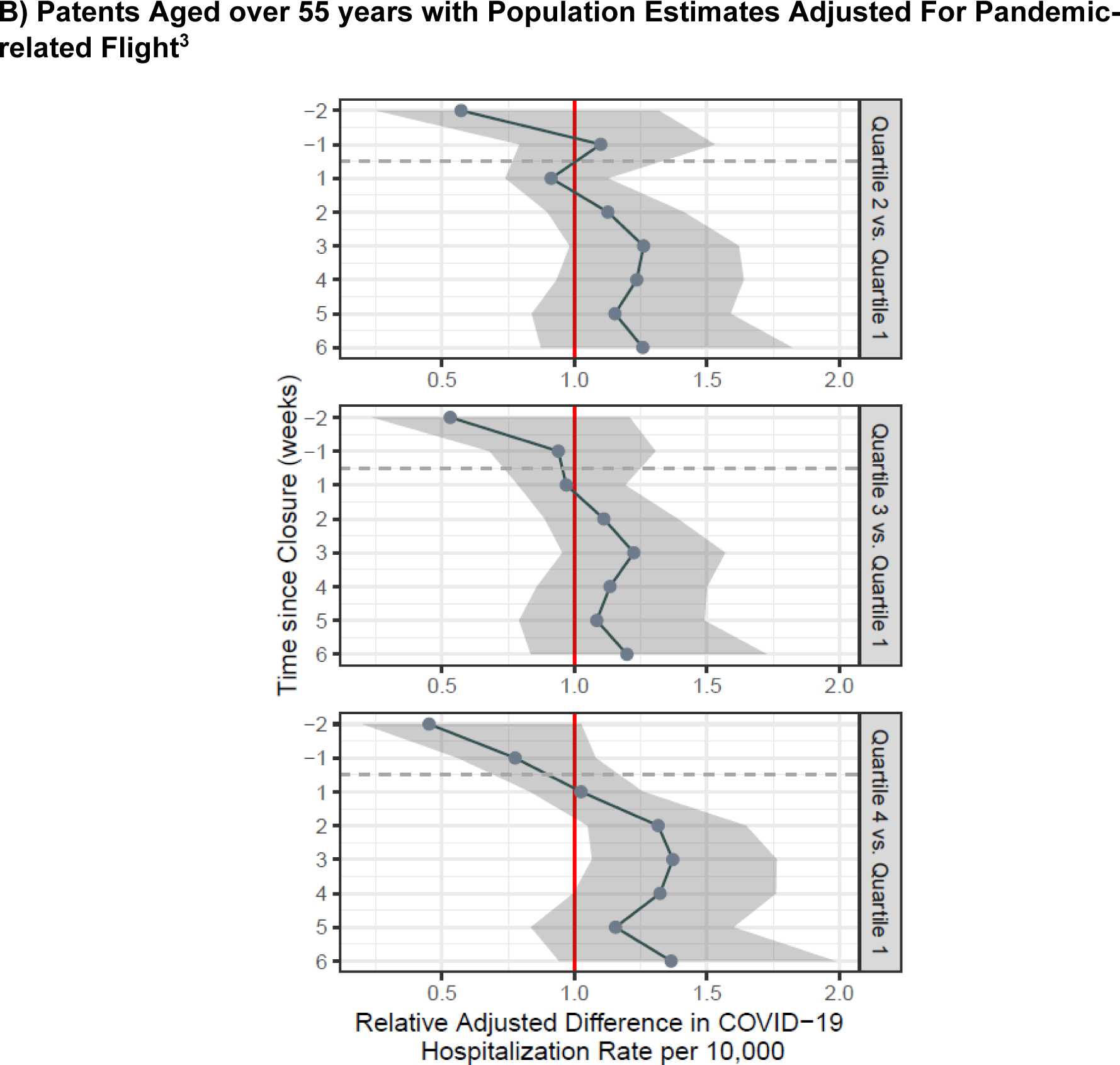
Adjusted Difference-in-Differences Estimates of the Association Between School Closure and Adjusted COVID-19 Hospitalization Rates by Quartiles of Multigenerational ZIP Codes, with Quartile 1 as reference.^1,2^. 1 In all panels, the Y-axis represents the number of weeks relative to the school closure order, with event weeks 2 weeks before the exposure, and up to 6 weeks after the exposure 2 Adjusted for the following ZCTA-level covariates: percentage of overcrowded households (defined as estimated number of housing units with more than one occupant per room, divided by the number of occupied housing units), percentage of White residents, percentage of residents living under the Federal Poverty Line, and median income in USD, 2018 – all taken from the American Community Survey 2018 5-year estimates. 3 ZCTA-level population shifts calculated from census tract estimates of population shifts between taken from publicly available cellular device data from Teralytics^1^ between January 1, 2020 and April 15, 2020, obtained from the New York Times.^2^

Figure 3B shows results that account for pandemic-related flight. Comparisons of adjusted COVID-19 hospitalization rates between Q1 and Q2, and Q1 and Q3 were not statistically significant throughout the study period. However, the relative difference in COVID-19 hospitalization rates between Q4 and Q1 increases towards a statistically significant difference three weeks after lockdown; at this time point, a 37% increase (95% CI: 1.07 – 1.76) in COVID-19 hospitalization rates is observed.

### Sensitivity Analysis

Estimates from the sensitivity analyses were largely consistent with those from the primary analysis (Supplement eFigure 3). When we considered multigenerational households and overcrowded households together in the same modelling strategy, we found that the association between city-wide lockdown, ZCTAs with higher proportions of multigenerational households, and COVID-19 hospitalization rates was attenuated, but remained significant (Supplement eFigure 4).

## Discussion

In this first empirical study examining the relationship between city-wide lockdown and COVID-19 hospitalization rates of older adults in multigenerational households, we found that the city-wide lockdown was associated with a transient and disproportionate increase in COVID-19 hospitalizations among individuals older than 55 years residing in ZCTAs with higher proportions of multigenerational households, after adjusting for socioeconomic characteristics. The estimated association was consistent with the transmission dynamics during the exponential growth phase of COVID-19 in NYC, specific to multigenerational households, and had its greatest effect three weeks after lockdown measures were undertaken. These results were attenuated by pandemic-related flight, but remained significant in the ZCTAs with the highest proportion of multigenerational households.

Our results add crucial ecological evidence to the emerging literature on household structure and COVID-19 risk. Moreover, they add empirical evidence to simulation studies that postulated deleterious effects of school closures - the first city-wide lockdown measure introduced.^18^ By assessing the effect of a city-wide lockdown with temporal and geographic specificity, our study contributes to this body of literature by illustrating how such blanket public health measures may contribute to uneven and transiently increased COVID-19 risk among older individuals residing in multigenerational households. Multigenerational households follow a socioeconomic gradient,^28^ and racial/ethnic differences between single and multi-generational households exist.^29,30^ The well-documented racial and ethnic disparities in COVID-19 related outcomes have been explained through mechanisms such as the disproportionate burden of disease among minority groups,^31,32^ limited economic opportunity,^33^ and other structural determinants.^34^ Our findings add another critical element to this discussion, introducing evidence for a link between multigenerational household structure and COVID-19 hospitalization. Our findings are in accord with various biological mechanisms of COVID-19 transmission such the relative differences in age-related outcomes from COVID-19, the role of proximity in viral transmission^2^ and subsequent severity of infection,^4^ and the 14-day incubation period (evidenced by the greatest difference in hospitalization rates being seen three weeks after lockdown measures).

Our findings should not be interpreted to suggest that NPIs such as city-wide lockdowns do not work. They clearly do. Instead, they suggest that the impact of such measures is not uniform across households and neighborhoods. Early reports in the pandemic underlined the age-dependent morbidity related to SARS-CoV2 infection.^35^ Therefore, it is logical that mandated home confinement including of those including school-age children during COVID-19’s exponential rise may disproportionately effect households where both young and older individuals reside.^36^ Our analysis does not establish causality, but it does suggest that NYC’s experience with its lockdown during the initial COVID-19 wave reflects this phenomenon. Identifying whether these findings are replicable in other cities, as well as assessing other potential secondary effects from different social distancing measures, remain an important area of future research.

### Limitations

This study has five limitations. First, despite attempts to address unobservable confounding influences, we cannot rule out the role of residual confounding due to the observational nature of the study. This is particularly relevant in the use of control covariates which acted as proxies for socioeconomic risk at ZCTA-level. Although we were careful to note which factors may act to mediate and confound the relationship under study, there could be other unmeasured time-varying factors. Second, our data were at the ZCTA level and did not allow us to evaluate individual COVID- 19 hospitalization risk. Third, our analysis does not quantify the relative COVID hospitalization risk of not undertaking city-wide lockdown on multigenerational households, which are likely to outweigh the impact of lockdown. Nonetheless, our results indicate that the risks and benefits of lockdown vary by different populations, with particularly stark consequences for multigenerational households. Fourth, hospitalization rates may not accurately reflect the remaining population base of ZCTAs in NYC because of pandemic-related flight, which favored wealthier neighborhoods.^25^ This may have reduced the total population at risk in our analysis, particularly in ZCTAs with lower proportions of multigenerational households, and led to overestimation of the neighborhood-level effects described. However, our use of mobile phone data attempted to address this issue. Lastly, we could not test the parallel trends assumption of our difference-in-differences design. However, pre-lockdown differences in adjusted COVID-19 hospitalization rates between quartiles were insignificant.

### Conclusions

In its first COVID-19 wave, NYC’s lockdown was associated with increases on COVID-19 hospitalization rates for individuals over 55 years of age in ZCTAs with higher proportions of multigenerational households. These findings highlight unanticipated interactions between structural factors such as housing and social distancing measures that may have contributed to greater transmission risks of select vulnerable groups in the pandemic.

## Data Availability

Data is available through a data use agreement with the NYC Department of Health and Mental Hygiene. Therefore, the authors are not able to share the raw data.

## Acknowledgements

We would like to acknowledge the New York City Department of Health and Mental Hygiene for access to data and Dr. Amelia Bond (Weill Cornell Medicine, Department of Population Health Sciences) for assistance in the use of methods.

