## Supplement for "Association between City-wide Lockdown and COVID-19 Hospitalization Rates in Multigenerational Households in New York City"

**Table of Contents**

eFigure 1: COVID-19 hospitalizations by ZIP Code Tabulation Area, cumulative from February 23 to May 23, 2020

eFigure 2: Model specification for primary analysis

eFigure 3: Difference-in-Differences Estimates of the Association Between School Closure and COVID-19 Hospitalization Rates by Quartiles of Multigenerational ZIP Codes and Adjusted for Multiple Socioeconomic Factors, with Quartile 1 as reference

eFigure 4: Difference-in-Differences Estimates of the Association Between School Closure and COVID-19 Hospitalization Rates by Quartiles of Multigenerational ZIP Codes and Adjusted for Multiple Socioeconomic Factors, with Quartile 1 as reference

**eFigure 1: Map of New York City with cumulative COVID-19 hospitalizations by ZIP Code Tabulation Area from February 23 to May 23, 2020**

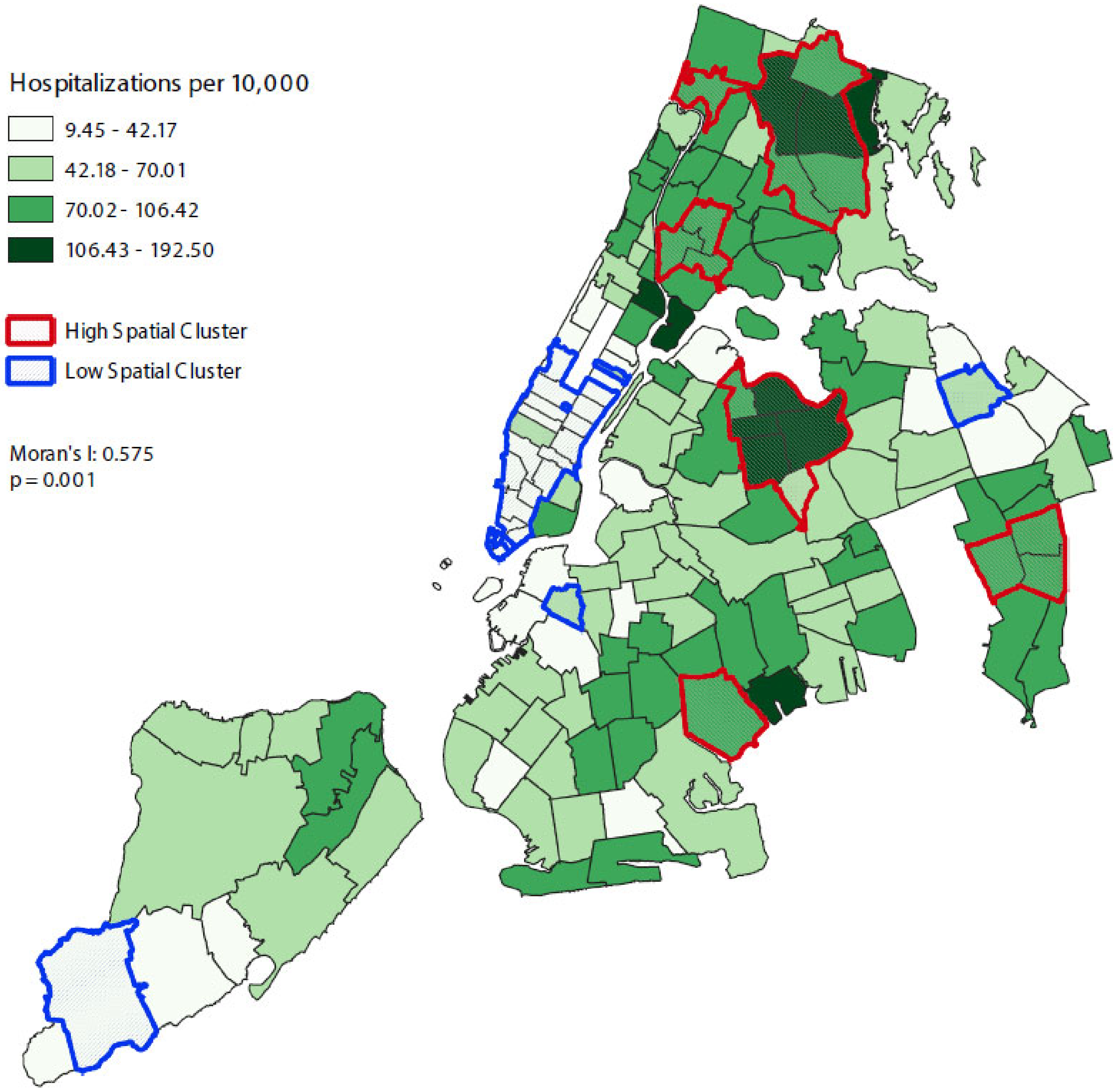

**eFigure 2: Model specification for primary analysis**

The generalized linear model we employed for the mean hospitalization count in each ZCTA in each week is of the following form:

$$\mathrm{E}\left(\log\left(10{,}000 \times \frac{Y_{it}}{\mathrm{Population}_i}\right)\right) = \beta_0 + \sum_{j=-2}^{6} \beta_{j,\mathrm{MG}_i}\left(\mathrm{Time}_j \times \mathrm{MG}_i\right) + \mu_i + \nu_i + \theta_t + \varepsilon_{ijt}$$

where:

- $Y_{it}$ denotes the number of hospitalizations in ZCTA $i$ in week $t$, where $t$ ranges from -2 (two weeks before school closure) to 6 (six weeks after school closure). The date of school closure is March 16, 2020. The $t$ = -2 count aggregates hospitalizations from the weeks of February 23, 2020 and March 1, 2020.
- $\mathrm{Time}_j$ is a binary variable indicating whether or not $j = t$. Time $t = 0$ (the week of lockdown and school closures) is treated as the reference level.
- $\mathrm{MG}_i$ is a categorical variable indicating which quartile of multigenerational housing ZCTA falls into (Q1, Q2, Q3, Q4). Quartile 1 is treated as the reference level.
- The quartile-specific coefficients $\beta_{j,\mathrm{MG}_i}$ are plotted in Figures 2 and 3. These coefficients can be interpreted as the difference in log hospitalization counts between quartile $\mathrm{MG}_i$ and quartile 1 at time $j$, relative to time 0. When exponentiated, they are interpreted as multiplicative factors rather than differences.
- $\mu_i$ denotes ZCTA-level fixed effects, including the effect of the ZCTA's median income, percentage of residents who are white, and percentage of residents below the federal poverty threshold.
- $\nu_i$ denotes a spatial random effect term for ZCTA $i$. In particular, it is a conditionally autoregressive term with a Besag-York-Mollié specification; this means that conditioned on the spatial random effect terms for all other ZCTAs, $\nu_i$ is modeled as normally-distributed with mean $\frac{1}{|N_i|}\sum_{j \in N_i} \nu_j$, where $N_i$ are the indices of the immediately-neighboring ZCTAs.
- $\theta_t$ denotes a weekly time fixed effect that captures trends in the outcome not explained by the other terms in the model.
- $\mathrm{Population}_i$ is the population of ZCTA $i$.

**eFigure 3: Difference-in-Differences Estimates of the Association Between School Closure and Adjusted COVID-19 Hospitalization Rates by Quartiles of Multigenerational ZIP Codes with Inclusion of all Socioeconomic and Clinical Risk Factors,[1] with Quartile 1 as Reference[2]**

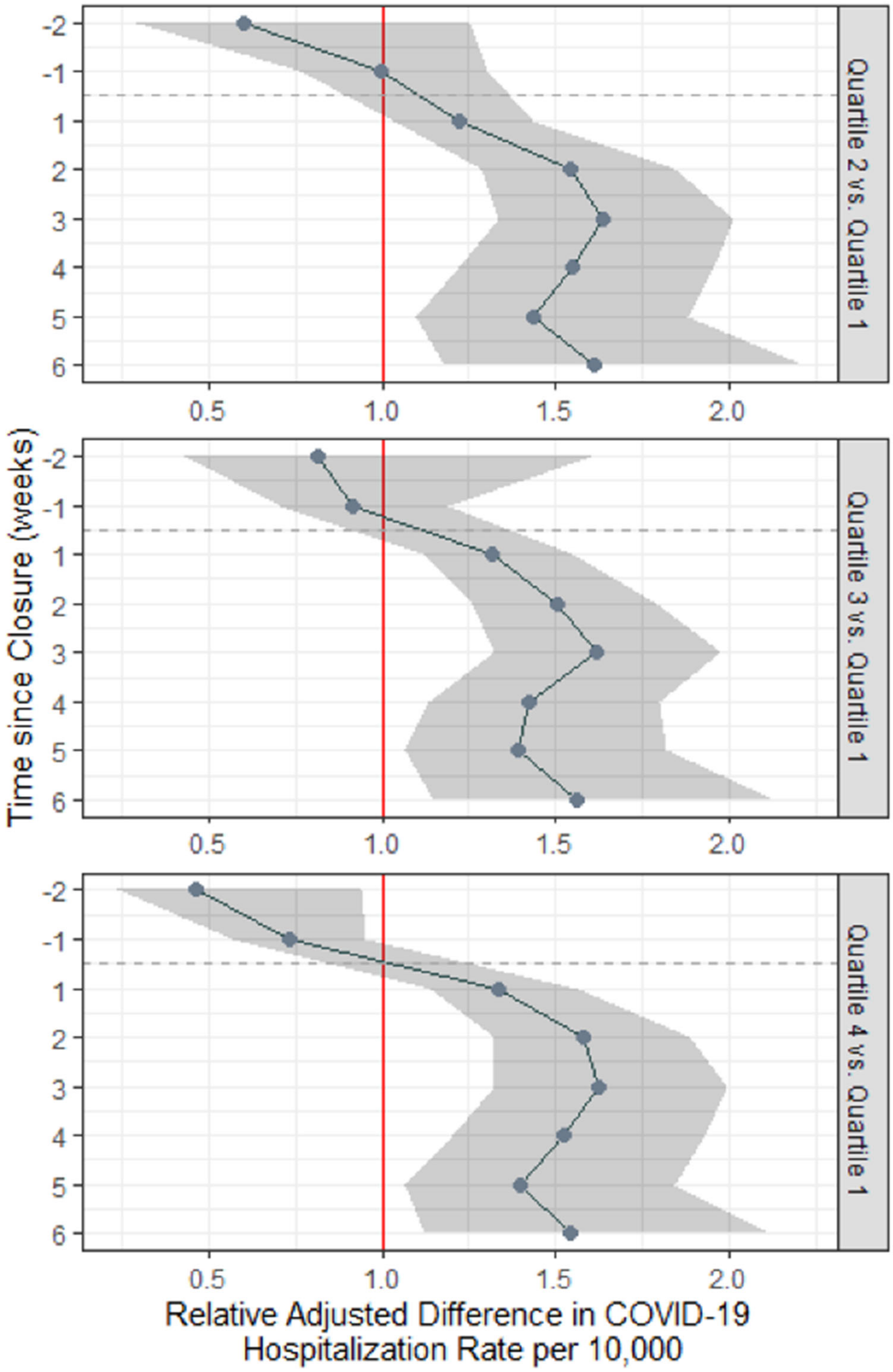


1 This model controlled for the following covariates: ZCTA-level prevalence (in percentage) of obese adults (defined as body mass index [BMI] ≥ 30 kg/m2), adults who smoke, and adults with coronary heart disease, hypertension, diabetes, asthma, or chronic obstructive pulmonary disease (COPD) – taken from the CDC 500 Cities Dataset; and additional socioeconomic factors: ZCTA-level estimates of total population, percentage of patients living below the federal poverty line (FPL), median income ,

percentage of White residents, and percentage of overcrowded households (defined as estimated number of housing units with more than one occupant per room, divided by the number of occupied housing units) - all taken from the ACS 5-year estimates 2018 - , and percentage of essential workers by ZCTA, identified from service-oriented non-public roles using Census Industrial Classification Codes in the following categories: 1) public transit workers; 2) grocery, convenience and drug store workers; 3) trucking, warehouse and postal service workers; 4) healthcare workers; 5) childcare, homeless, food and family service workers; and 6) building cleaning service workers – we replicated the same methodology as employed by the New York City Office of the Comptroller (Scott S [2020] New York City's Frontline Workers. New York City: City of New York, Office of the Comptroller).

2 All COVID-19 hospitalizations included in this analysis

**eFigure 4: Difference-in-Differences Estimates of the Association Between School Closure and Adjusted COVID-19 Hospitalization Rates by Quartiles of Multigenerational ZIP Codes with the inclusion of Quartiles of Overcrowded ZIP codes[1], with Quartile 1 as reference[2]**

**a) Quartiles of Multigenerational ZIP codes**

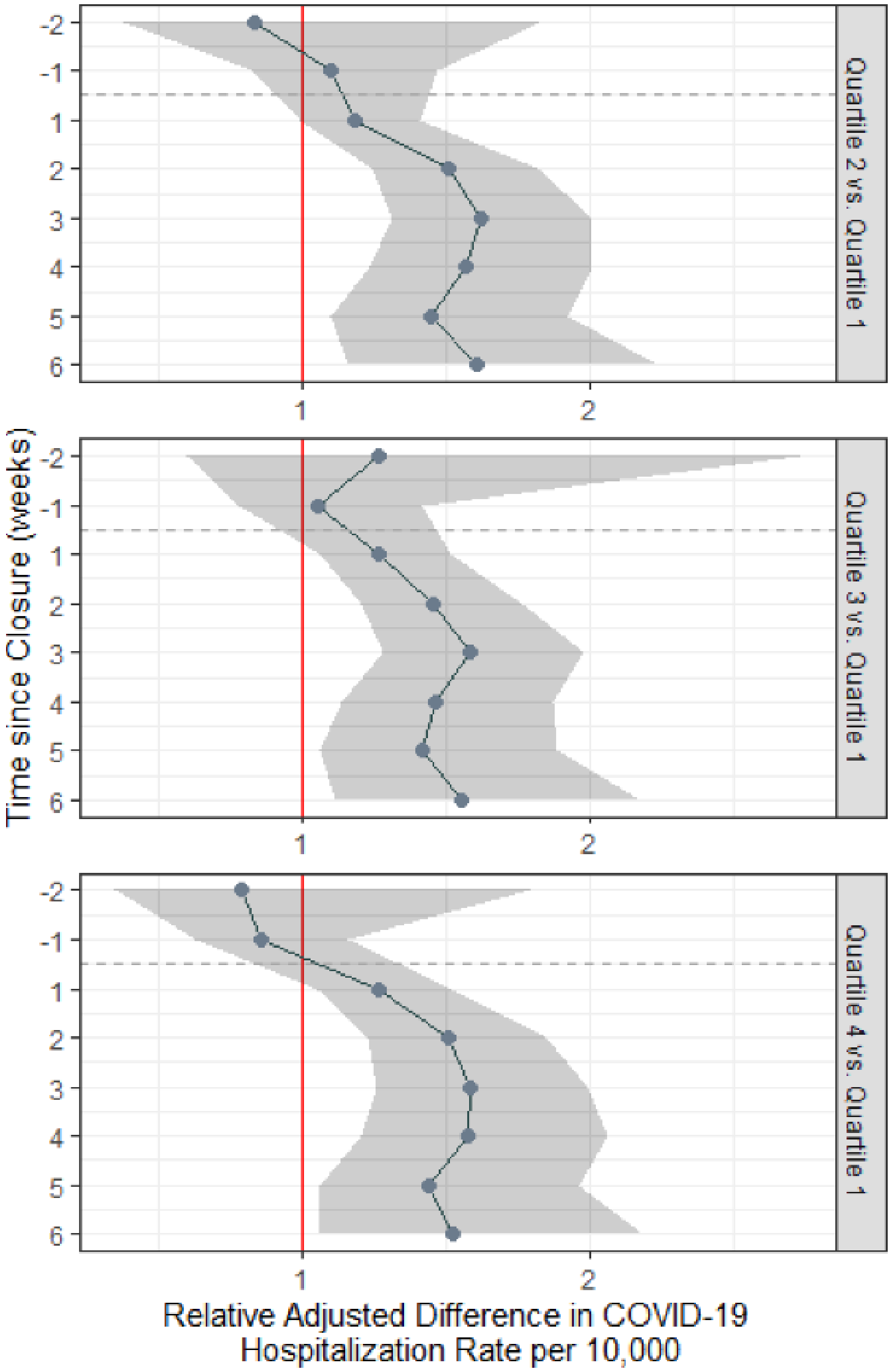

**3b) Quartiles of Overcrowded ZIP codes[3]**

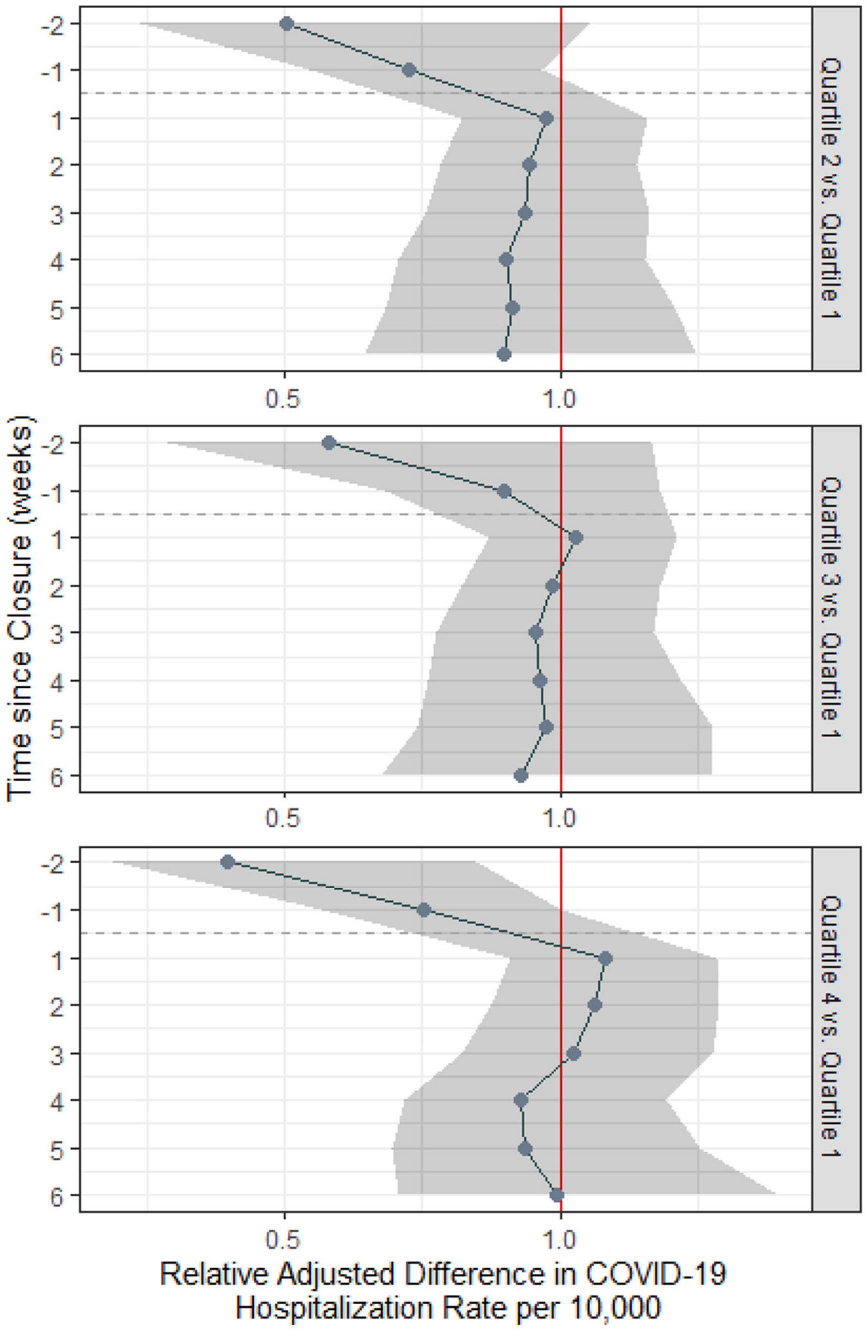


1 Models in A) and B) controlled for the following covariates: percentage of patients living below the federal poverty line (FPL), median income in 2018 USD, percentage of overcrowded households (defined as estimated number of housing units with more than one occupant per room, divided by the number of occupied housing units) and percentage of White residents (all taken from the ACS 5-year estimates 2018

2 All COVID-19 hospitalizations included in this analysis

3 Represents the coefficients and 95% Bayesian credible intervals of the interaction between time in weeks (indexed at t = 0 for the week of school closure) and quartiles of ZCTAs with overcrowded

households, after accounting for the interaction effect between time and quartiles of ZCTAs with multigenerational households.
